# Dynamic Prediction of Mortality Risk Following Allogeneic Hematopoietic Stem Cell Transplantation

**DOI:** 10.1101/2025.02.17.25322391

**Authors:** Philipp Spohr, Rebecca C. Fröhlich, Sebastian Scharf, Anna Rommerskirchen, Jonathan Bobak, Sarah Schweier, Paul Jäger, Guido Kobbe, Sascha Dietrich, Alexander T. Dilthey, Birgit Henrich, Klaus Pfeffer, Rainer Haas, Gunnar W. Klau

**Author notes:** Shared First Author / Shared Last Author (Philipp Spohr, Rebecca C. Fröhlich, Klaus Pfeffer, Rainer Haas, Gunnar W. Klau).

## Abstract

Allogeneic hematopoietic stem cell transplantation (alloHSCT) is a potentially curative treatment for high-risk hematological malignancies, but early mortality within 100 days remains a significant challenge, affecting approximately 18% of patients. Identifying high-risk patients early is critical for timely interventions. While static risk scores like the Hematopoietic Cell Transplantation-Specific Comorbidity Index (HCT-CI) and Endothelial Activation and Stress Index (EASIX) are valuable, they do not adapt to patients’ clinical trajectories. Leveraging the increasing availability of digital medical datasets, we developed a dynamic risk stratification system based on a random forest approach to continuously predict mortality risk from day 0 to day 30 post-transplantation.

Analyzing data from 847 patients undergoing alloHSCT between 2004 and 2019, our model achieved high classification performance, with AUROC scores continuously exceeding 0.8 from day 17, and effectively stratified patients into high, moderate, and low-risk groups. Feature importance analysis revealed a shift over time, with early predictions utilizing diverse variables and later stages relying more on inflammation-related bloodwork parameters. This dynamic approach surpasses static methods, demonstrating improved predictive performance and adaptability across treatment phases. To promote reproducibility and broader application, we provide open access to the software and accompanying bloodwork time-series dataset.

## Introduction

Allogeneic hematopoietic stem cell transplantation (alloHSCT) is an efficient treatment for patients with high-risk hematological malignancies and for many of them the only option with curative potential. While the overall mortality of alloHSCT has steadily decreased due to reduced intensity conditioning regimens and advances in supportive therapy, a significant number of patients (approx. 18%) still die early within 100 days after transplantation, due to early relapse, infections, or graft-versus-host disease (GvHD) [1]. The 100 day mortality is a common indicator for the combined risk a patient is exposed to.

Identifying high risk patients is crucial to provide early extended diagnostic procedures and therapeutic intervention. Clinical risk scores such as the Hematopoietic Cell Transplantation-Specific Comorbidity Index (HCT-CI) [2] or the Endothelial Activation and Stress Index (EASIX) [3] are valuable predictors for mortality risk, however, without taking into account the patient’s clinical course, as they are typically determined only at admission and not continuously updated.

The increasing digitalization of medical datasets allows the utilization of a wide set of parameters and therefore enables the use of machine learning-based algorithms for risk prediction. Although final therapeutic decisions remain with physicians, such systems can be of valuable assistance. For their acceptance, explainability remains an important factor. While approaches based on complex machine-learning algorithms such as convolutional neural networks (CNNs) show promising results [4], the interpretability of their predictions is difficult. In contrast, tree-based models naturally facilitate transparency by providing interpretable feature importances. Shouval et al. were able to train a decision tree-based model to predict 100 day mortality after alloHSCT that achieved an AUROC score of 0.701 using only 20 variables [5]. Arai et al. achieved an AUROC score of 0.622 predicting acute GvHD using an alternating decision tree implementation [6]. Nevertheless, both models only provide a prediction at the time of admission that is not updated throughout the treatment. Models that are continuously updated have also been developed, specifically for high-frequency data ([7], [8]) but remain rare for more sparse data and are also challenging to easily evaluate by clinicians. Thus, a dynamic risk stratification utilizing clinical and laboratory data sets would be a significant step forward to make predictions accessible.

Based on a group of 847 patients who underwent allogeneic HSCT between 2004 and 2019 at a single center, we present a risk stratification system that continuously evaluates each patient’s mortality risk from day 0 to day 30. Our results show that the system achieves a high classification performance (AUROC scores continuously over 0.8 starting at day 17) and accurately stratifies patients into three risk groups: high, moderate, and low. We also demonstrate the method’s increasing performance over time and observe a shift in feature importance. Early predictions utilize a broad range of features, while predictions between days 7–21 are mainly based on inflammation-related bloodwork parameters. This dynamic adaptation underscores the ability of our method to adjust its focus based on the patient’s actual clinical course.

## Results and Discussion

### A method for dynamic risk stratification

We present a machine learning-based risk assessment system that utilizes time-series data of bloodwork in addition to static parameters such as primary diagnosis or age to continuously evaluate an individual risk for each patient per day, starting from the day of transplantation as day 0 until day 30. Our system was trained and evaluated using a group of 847 patients receiving an allogeneic HSCT in the Department of Hematology, Oncology and Clinical Immunology of the University Hospital Düsseldorf between 2004 and 2019 (See Figure 1). A random forest model classifying a patient as either “deceased within 100 days” or “alive after 100 days” was trained separately for every prediction day on the corresponding feature matrix and a subsequent risk stratification strategy assigns each patient to “low”, “moderate” or “high” risk so that physicians can be continuously provided with a risk assessment increasing their alertness for complications and permitting adjustments of the current therapy.

**Figure 1:**
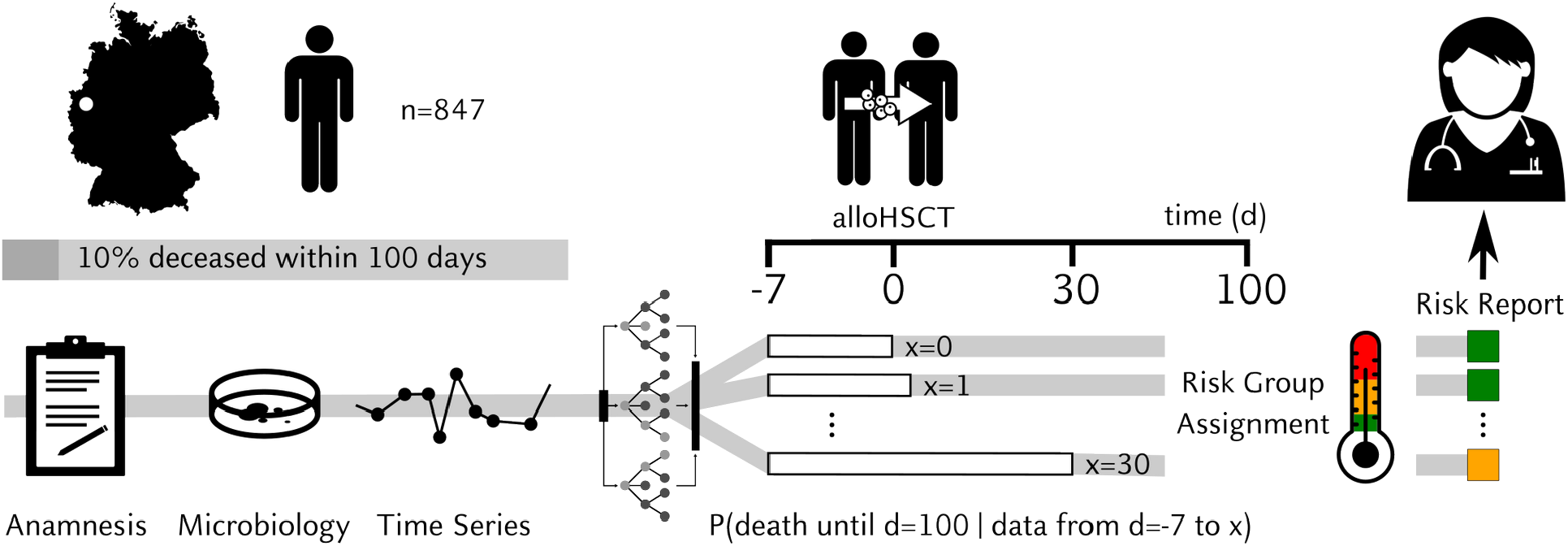
Overview of the study and method. Our cohort consists of 847 patients from the Department of Hematology, Oncology and Clinical Immunology of the University Hospital Düsseldorf who received an alloHSCT. Of those, 85 died within 100 days of transplantation. Our machine learning model predicts the patient’s risk of dying within 100 days, starting from the day of transplantation up to 30 days afterwards. The data consist of static anamnesis parameters such as diagnosis, age/sex; microbiology data such as detected pathogens/resistance testing as well as continually measured parameters such as hemoglobin, blood cell count and biochemical parameters starting from day -7 to the day of prediction. In addition, derived features describing the series of aforementioned parameters are calculated. We utilize a random forest approach: Multiple decision trees assess the risk independently and vote for a decision. Finally, we translate the risk probability into a discrete risk classification, labelling each patient on every prediction day as either “low”, “moderate” or “high” risk, thus providing easily parsable live assistance to clinicians.

For a better understanding we publicly share the software alongside with the comprehensive dataset (static parameters: anamnesis, diagnosis, age, sex; dynamic parameters: microbiology data [pathogens/resistance testing]; bloodwork and biochemical parameters starting from day - 7) for all patients undergoing allogeneic hematopoietic stem cell transplantation (alloHSCT), enabling full reproducibility of our work and its application to other cohorts (See Materials and Methods).

### Group assignment reliable for short-term mortality

The assignment to the risk group showed good overall performance with deceased patients frequently getting assigned to high risk and likewise surviving patients getting assigned predominantly to low risk. Analyzing risk group assignments for the test set of 253 patients (24 deceased and 229 alive beyond day 100 post-transplantation), we found that,over the entire observation period of 30 days, on average 45.93% of deceased patients were classified as high-risk in comparison to only 12.36% within the surviving patients. On the other hand, an average of 63.12% of the surviving group were classified as low-risk compared to only 29.83% in the group of deceased patients (See Figure 2, Panel A). In addition, the patients classified as low-risk within the group of deceased patients died primarily between days 76–100, suggesting better model performance for adverse events early after transplantation (See Figure 2, Panel B). It was interesting to note a general increase in the proportion of high-risk patients in both, the surviving and deceased patients, up to day ten, which relates to the most critical period of severe neutropenia (which is considered a strong risk factor for bacterial and invasive fungal infection [9]). Thereafter, we observe a decreased risk in the surviving group of patients with none of them remaining in the high-risk category on day 16. Different from that pattern, the deceased patients also show a tendency towards a lower risk, but during the following three days their proportion within the high-risk group is sharply raised to approximately 70%. This is of particular relevance as around day 17 the majority of patients should have a hematological (neutrophil) recovery which is associated with a decreased risk of complications [10]. The persistence of a patient in the high-risk group beyond day 17 is therefore a strong indicator and alarming sign that the mortality risk for the patient is high and potentially related to organ complications not necessarily related to the immunocompromised status. In general, patient risk trajectories were mostly constant progressions across risk levels without abrupt shifts. Over time, the moderate-risk group shrinks which reflects an increasing model certainty. Notably, individual misclassifications such as some high-risk patients surviving beyond 100 days still represented the status of patients well (See “Misclassifications reflect clinical complexity”). Supplementary Figure 1 illustrates all individual patient trajectories.

**Figure 2:**
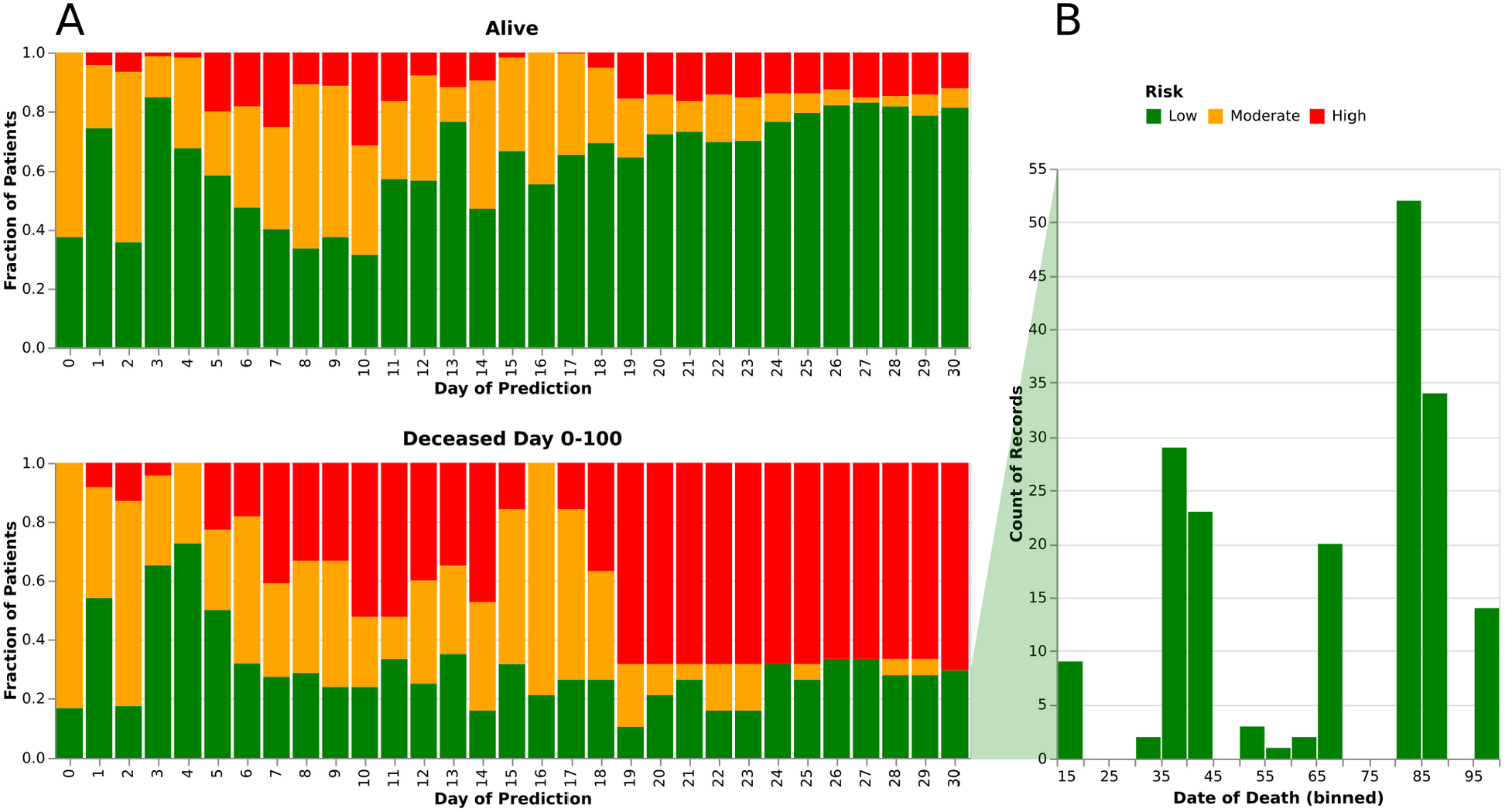
Overview of the risk group assignments for the test data set. A) The fraction of patients assigned to a specific risk group is shown for each prediction day separately for the two outcomes. B) The time point of death after alloHSCT (day 0) is shown for all “low-risk” predictions made for deceased patients.

### Model predictive performance increases with time

**Figure 3:**
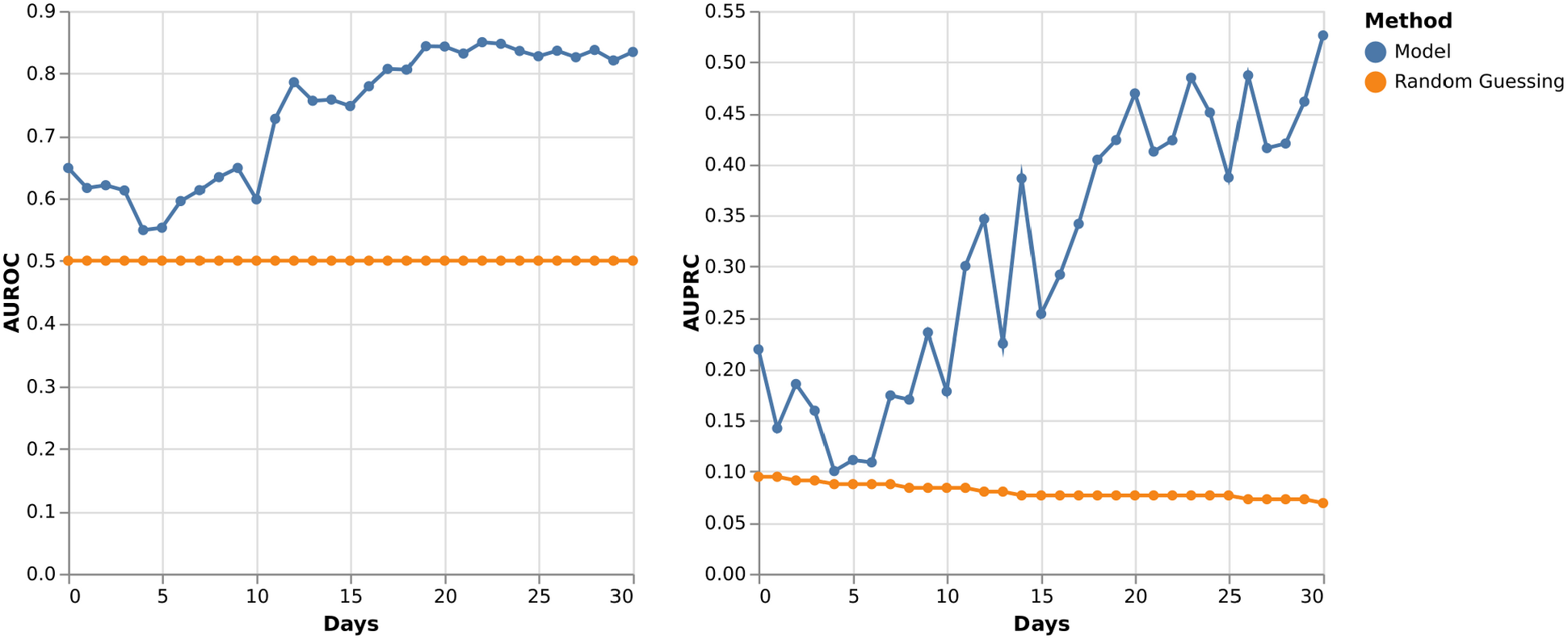
Overview of the model performance for individual days. Shown are (left panel) the Area under the Receiver Operator Curve (True Positive Rate vs. False Positive Rate) as well as (right panel) the Area under the Precision-Recall Curve for each prediction day. As a baseline we also plotted the performance of random guessing (assigning a random score between 0 and 1 to each row): For the AUROC this value is a constant 0.5 since FPR and TPR are symmetrical for random guessing; For the AUPRC the random guessing value depends on the class weights since the precision is affected by it.

As a basis for risk prediction, we trained a random forest model to predict 100-day mortality. Our model reaches a mean AUROC score of 0.74 (Std. Deviation 0.10, Min. 0.55, Max 0.85) and a mean AUPRC score of 0.31 (Std. Deviation 0.13, Min. 0.10, Max 0.53). We observed a steep increase in prediction quality starting at around day 10. The overall lower stability of the AUPRC is the result of an overall small test set size and class imbalance; in the test set only 9.8% (Day 0) - 6.9% (Day 30) of patients died. Notably, the model achieves an AUROC score of 0.648 for day 0 indicating an already usable risk prediction on the day of transplantation.

### Feature importance shifts throughout treatment course

**Figure 4:**
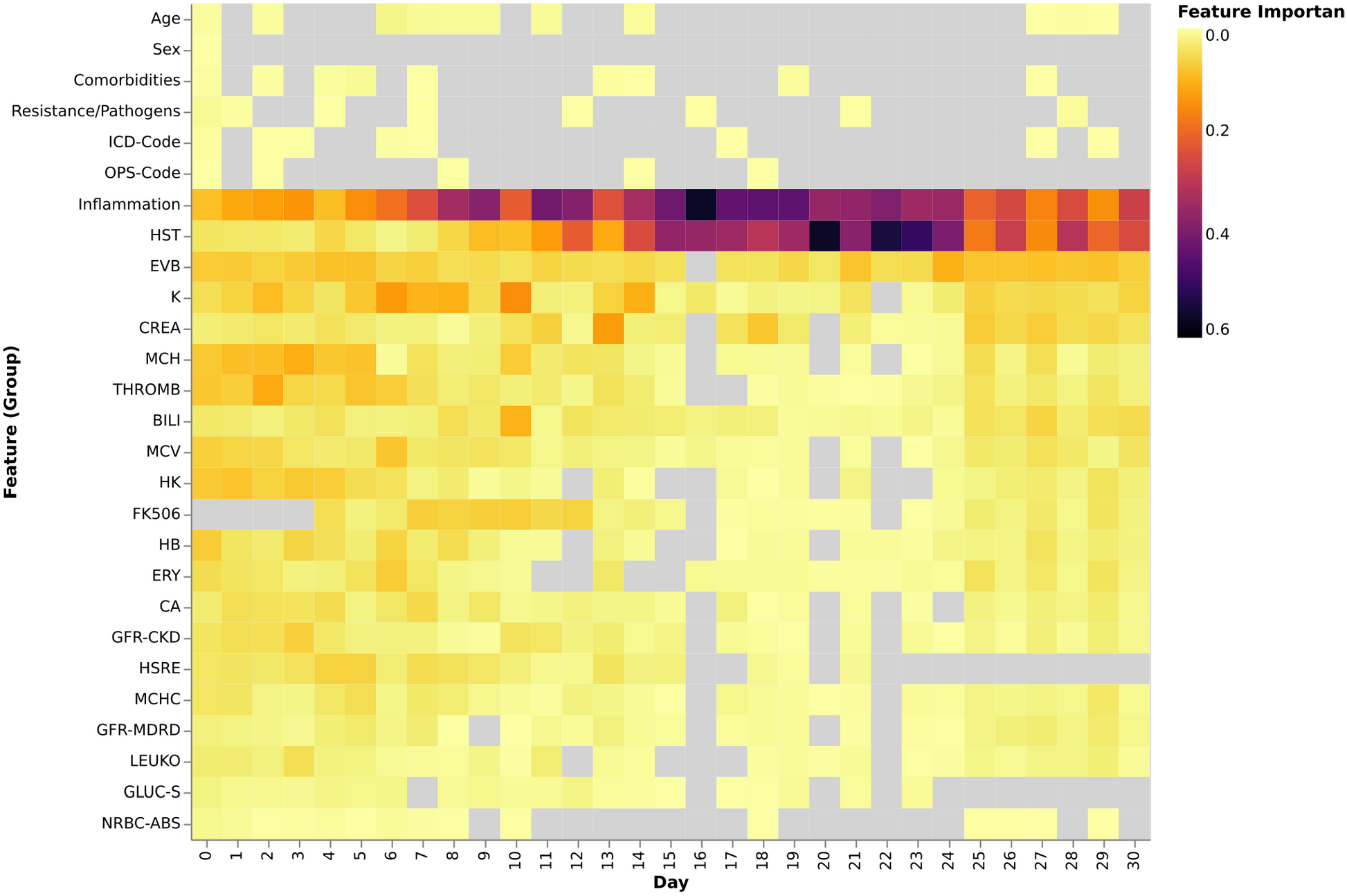
Overview of the feature importances. Features are grouped and their cumulative feature importance (Gini impurity decrease) is encoded via color. Age is a single feature describing the age at transplantation; Sex describes the biological sex of the patient; Comorbidities aggregates all known comorbidities prior to prediction; Resistance/Pathogens contains microbiological pathogen detections and resistance testing; ICD-Code contains primary diagnosis information; OPS-Code contains information about the transplantation; Inflammation contains c-reactive protein and interleukin 6; HST urea; EVB red blood cell distribution width; K potassium; CREA creatinine in serum; MCH mean corpuscular haemoglobin; THROMB thrombocytes; BILI bilirubin; MCV mean corpuscular volume; HK haematocrit; FK506 tacrolimus; HB haemoglobin; ERY erythrocytes; CA calcium; GFR-CKD glomerular filtration rate (CKD); HSRE uric acid; MCHC mean corpuscular haemoglobin concentration; GFR-MDRD glomerular filtration rate (MDRD); LEUKO leukocytes; GLUC-S glucose in serum; NRBC-ABS nucleated red blood cells; See “Data Preprocessing and Feature Engineering” for a detailed overview of all features. Features that are not used on a specific day for prediction are greyed out: This might be because 1) the feature was eliminated during feature selection, 2) the measurement was not available on that specific day or 3) the feature simply was not utilized by any decision tree.

The feature importance analysis highlights a strong emphasis on bloodwork values, particularly C reactive protein (CRP) and urea (HST), suggesting a correlation with inflammatory, renal function and protein catabolism. Variables such as age, sex, and other static parameters mostly achieve relevant feature importance values early on and late, whereas the bloodwork values dominate between days 7-24, indicating that the method prioritizes the patient’s current status rather than static characteristics in this “critical phase”. In general, we observed that the model gives measurements closer to the prediction day a higher importance (See Supplementary Figure 2). We noted that the resistance/pathogen features achieved non-zero importances and might therefore be worth exploring in risk assessment systems.

### Misclassifications reflect clinical complexity

**Figure 5:**
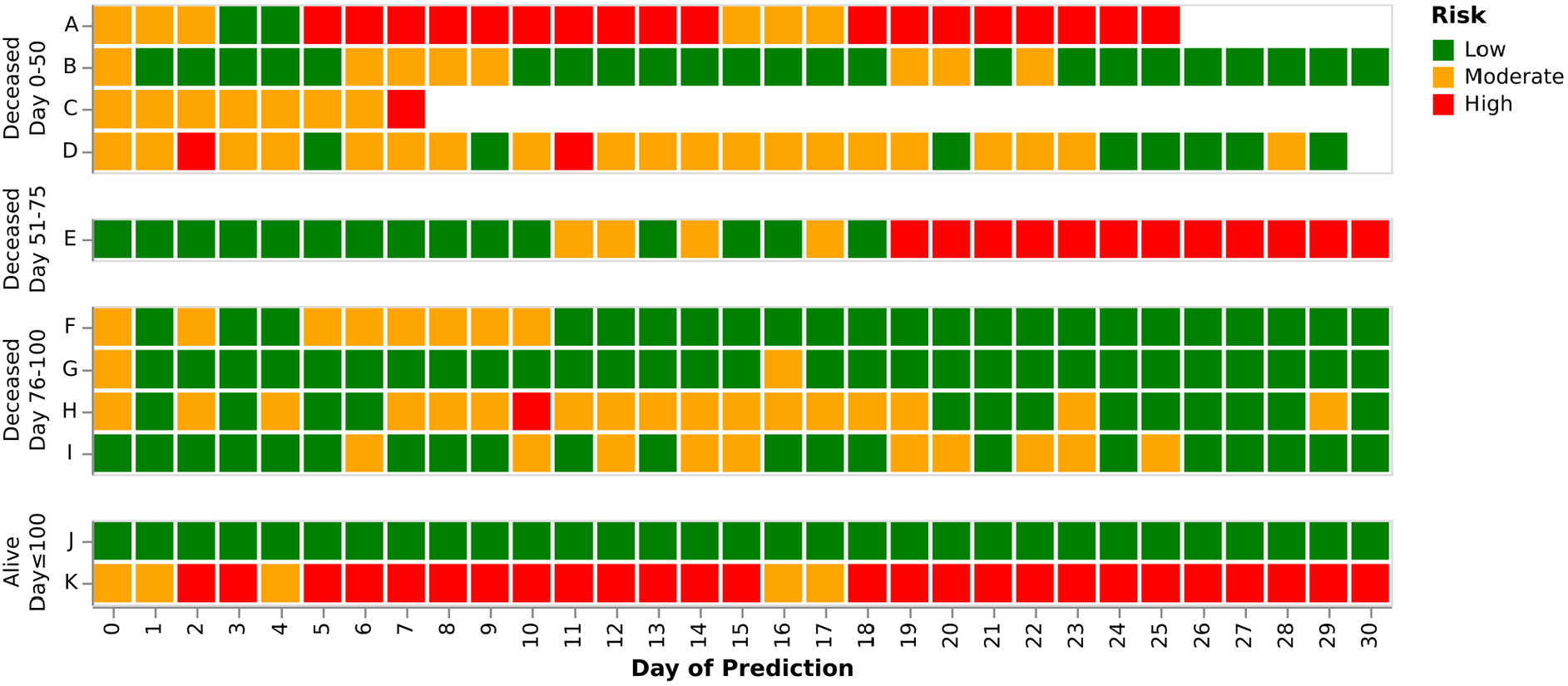
Exemplary risk prediction trajectories for a set of patients (with misclassifications overrepresented). **Patients A-K** are grouped by their outcome (day of death / or no death within 100 days) and their risk assignment is shown for every prediction day. “White” missing squares indicate that the patient will be deceased within 5 days.

The model predictions generally align well with clinical risk assessments of physicians, while there are certainly mismatches in some patients, reflecting the multifactorial complexity of patient outcomes. For instance, there are six patients (B, D, F, G, H, I) predicted to be low risk (green) who died post-transplantation of complications with “sudden onset” such as sepsis, multi-organ failure, or pulmonary embolism. Another example is patient F who was discharged in good health on day 21 following allogeneic HSCT. The patient developed a lethal acute liver and kidney failure without leading anteceding symptoms resulting in death on day 84. Similarly, patient I succumbed to a sudden cardiac death following a lung embolism.

On the other hand, there are examples where the model predicted high risk (red), while the patients survived. For instance, patient K was correctly classified as high-risk because of severe complications like invasive aspergillosis and acute GvHD. Fortunately, the patient survived, reminding us that probability assessments do not permit to forcast individual fates. We also found that predictions are more meaningful when patients are inpatients and thus clinical and laboratory parameters obtained regularly. For example, Patient D was followed as an outpatient between days 19 and 28 resulting in fewer measurements during this period. Clinical annotations for patients A-K are provided in Supplementary Note 1. Supplementary Figure 1 illustrates all individual patient trajectories. See Supplementary Note 2 for a mapping of letters to patient identification numbers used in the provided dataset.

### High risk predictions can precede risk recognition by clinicians

Determining whether the risk prediction system can assist the judgment of experienced clinicians and is able to predict critical cases earlier remains a challenging task. To address this issue we used the judgment of our author RH, an experienced clinician who manually examined the features for 19 deceased patients of the test who were classified as high-risk by the algorithm at some point post-transplantation (See Supplementary Note 2 for a mapping to patient IDs used in the provided dataset as well as the full clinical assessment). Thereby, we identified patients for whom the system provided added value for the clinical reasoning.

For instance, the model predicted Patient X (deceased on day 19) to be at high risk for the first time on day 8. While both the algorithm and clinical assessment eventually considered the patient as high risk, this classification would not have been made that early based solely on clinical judgment since the sole results of the bloodwork did not show any obvious increased risk in comparison to the anteceding days.

Similarly, Patient Y was first flagged as high risk on day 7 and died on day 39. The shift in risk assessment from moderate to low risk occurring from day 1 to day 2 appeared implausible; but the upgrade from day 4 to day 5 to moderate risk would probably not have been made by clinicians. The same is true for the upgrade to high risk, as in the light of stable CRP values clinicians might have underestimated the continuous rise in urea levels observed since day 4. The course of this patient might serve as an example for the utility of the algorithm in recognizing complex patterns with subtle changes that may be missed by purely human reasoning.

Far from being a state of the art prospective comparison, these examples show the value of algorithmic systems to complement clinical decision-making. In total, we found that for 11 of the patients the algorithm made a high-risk classification earlier than the clinician would have done. On the other hand, for five patients and 3 patients the clinician’s upgrade to high risk was somehow earlier or at the same time, respectively

### Additional and improved data sources hold large potential

The next step could be an evaluation of our findings on an independent dataset of alloHSCT patients from other centers to see whether our AI model is of general value, while a prospective study where physicians assign patients to risk groups to compare their assessments to our model’s predictions is apparently not feasible taking into account the great number of patients needed to be recruited within a reasonable time.

With all these caveats and shortcomings of a retrospective study in mind we believe that our results are at least a proof-of-principle for the utility of real time AI based predictions to assist the physicians in their day to day clinical reasoning. Further refinements of the system can be achieved by incorporating more parameters and features such as time from diagnosis to transplantation, the number of prior transplantations, the graft source, and previous iron chelation therapy could be included [11]. This process could be further fostered by the growing digitalization of the patients data including vital parameters or the results of imaging examinations such as CT or MR scans.

## Materials and Methods

### Software and data availability

The software and data are available as open source on github: **https://doi.org/10.5281/zenodo.14779813**. The software makes use of scikit-learn [12] and tsflex [13] and is implemented as a Snakemake [14] workflow, allowing ease-of-use and scalability to large datasets and cloud computing environments.

### Patient cohort and characteristics

**Figure 6:**
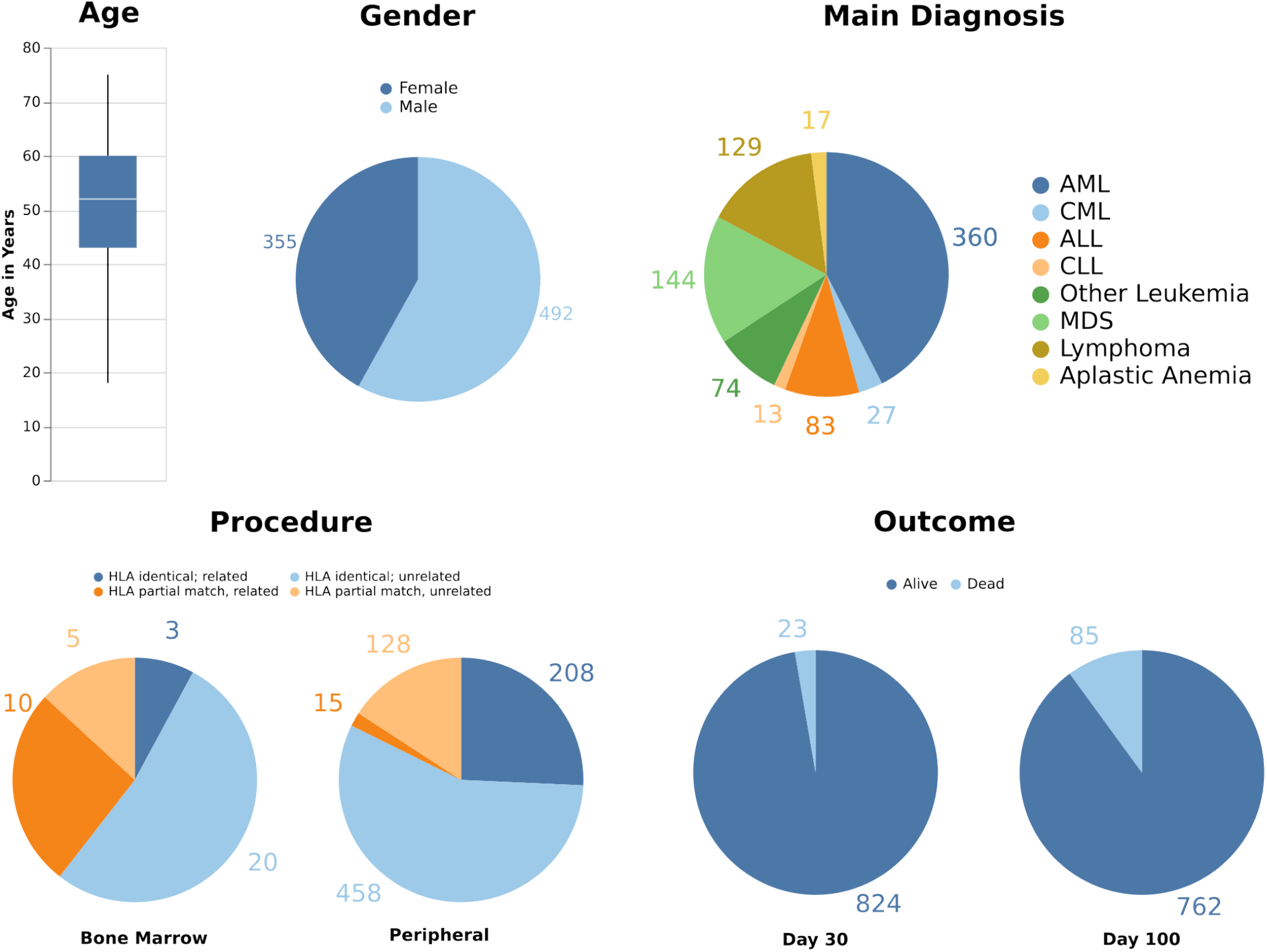
Overview of the main cohort from the University Hospital Düsseldorf and its characteristics.

The main cohort was drawn from cases admitted to the UKD for alloHSCT in the years from 2004 until 2019. For a detailed overview of the characteristics and the specific ICD-10 GM codes refer to Supplementary Table 1.

### Data preprocessing and feature engineering

The raw data extracted from the system is transformed into one matrix per prediction day where every row is a case and every column is a feature. The following features are generated:

#### Demographic Features

Sex is encoded as a binary feature, Age in days is encoded as an integer

#### Comorbidities

The risk groups are modeled after the HCT-CI, namely: Arrhythmia, Heart disease, Chronic inflammatory bowel diseases, Diabetes, Cerebrovascular diseases, Psychiatric disorders, Liver dysfunctions, Obesity, Infections, Rheumatic diseases, Gastric ulcer, Kidney dysfunctions, Lung dysfunctions, Tumor. For each group, the amount of associated diagnoses documented for a stay with a discharge date within 365 days prior to transplantation is encoded as an integer. See Supplementary Note 3 for a mapping of ICD-10-GM codes to the groups.

#### Resistances & Pathogen Detection

Microbiome resistance testing is encoded as a binary feature for each combination of pathogen, drug and both aerobic and anaerobic blood cultures. Three additional binary feature columns are generated for the confirmed detection of MRSA, VRE, and MRGN within a year prior to the HSCT respectively.

#### ICD Codes and OPS Codes

Every OPS^1^ Code (encoding donor/receiver match, blood vs. peripheral, and HLA match) and every main diagnosis ICD-10-GM^2^ Code in the dataset is represented as a binary feature (meaning the patient has the code documented in the hospital stay across transplantation as a primary diagnosis).

#### Blood Work Measurements and Time Series

We extracted all clinical blood work parameters starting from seven days before transplantation up to the day of prediction from the laboratory information system. Blood values are filtered if the median number of measurements across all measured patients is below 80% of all days between -7 and prediction. If multiple measurements exist on the same day the median is chosen, missing values are replaced with a constant value of -1000. For the values we calculate derivative time series features as feature columns in addition to the raw measurements, refer to Supplementary Note 4 for a full overview and explanations.

The time series features were calculated using tsflex which allows gaps within the time series; thus, the constant values of -1000 are only used for the raw measurement column.

Rows are only labeled as deceased within 100 days if a clear death event was recorded, and only labeled as alive if data indicating that is available after 100 days, otherwise the row is treated as “lost to followup” and removed.

Rows that contain a death within 5 days after prediction are also removed to avoid prediction of imminent events (Note that this leads to a decrease of 33 rows over time).

### Model training and evaluation

A random forest model classifying a patient as either “deceased within 100 days” or “alive after 100 days” is trained separately for every prediction day on the corresponding feature matrix. Since the number of rows decreases over time we aim to have a consistent test and training set (and thus ensuring comparability) for all prediction days by performing a stratified split on all rows that are contained on every prediction day into 30% test data and 70% training data. The remaining 28 rows - representing patients deceased within 35 days - are then successively assigned to the sets such that for every day the split ratio stays as close to 30/70 as possible.

To optimize the hyperparameters for each method, five-fold stratified cross validation with five repeats is used in combination with a full grid search: For every possible hyperparameter choice the training set is split five times into five disjunct folds, four of which are used to train the model and one of which is used for evaluation. In the end, the hyperparameter choice resulting in the highest average F1 score is used to train the model on the entire training data and then evaluate it on the test data.

### Hyperparameter settings

For our random forest model we make use of trees with depth 2. For every decision in the tree we check all possible splits and take the one resulting in the highest impurity decrease. The other parameters were determined during the training using a full grid search with the following options (where m is the total number of available features):

Number of features per tree (maxFeatures): log2(m), sqrt(m)

Minimum amount of samples per leaf (minSamplesLeaf): 2, 4, 7

Number of decision trees (n_estimators): 100, 150, 200, 250

Target number of features for the feature selection: 20, 40, 100, 200, m

Refer to Supplementary Table 2 for the chosen hyperparameters for each prediction day.

### Feature importance and selection

Feature selection is performed based on mutual information.

Reported feature importances were calculated using the rf.feature_importances_ function from scikit (mean of the Gini-impurity decrease across all trees).

### Risk group assignment

To determine the assignments of patients to either the “low”, “moderate” or “high” risk groups on a specific day two thresholds *θ*_*H*_ and *θ*_*L*_ on the predicted scores are used. A patient achieving a prediction score of *θ*≥ *θ*_*H*_ is assigned to the high risk group, a patient achieving a score of *θ*≤*θ*_*L*_ is assigned to the low risk group. Remaining patients are assigned to the moderate risk group.

Using the training dataset, the threshold *θ*_*H*_ is chosen such that at most 20% of non-deceased patients fall into the high risk group. Analogously, the threshold *θ*_*L*_ is chosen such that at most 5% of deceased patients fall into the low risk group. In other words, we control the False Positive Rate to be less than 20% and the False Negative Rate to be less than 5%.

## Data Availability

All data produced in the present work are contained in the manuscript

https://doi.org/10.5281/zenodo.14779813

## Ethics

The study was approved by the “Ethikkommission an der Med. Fakultät der HHU Düsseldorf” (Study-Nr.: 2019-513).

## Acknowledgements

We want to express our gratitude to the nursing and medical staff of the UKD for supporting our work, collecting samples and data and providing excellent care to the patients. For providing excellent technical assistance we want to thank Max Jakub Ried.

## Author Contributions

RF developed and evaluated the software. PS supervised the software development and evaluation. PS, RF and SeS wrote the initial manuscript draft; GKl and RH supervised manuscript preparation and performed editorial revisions. PS, JB and GKl supervised the model development. SaS wrote software for data extraction from medical databases. SeS and PS programmed additional software for data extraction and processing of clinical records. RH and SD provided a detailed clinical perspective. AR coordinated ethics. PJ and GKo coordinated data acquisition. AD, BH, KP, GKl and RH initiated and supervised the project. All authors have contributed to the internal review process, read and approved the final manuscript.

## Footnotes

1 https://www.bfarm.de/EN/Code-systems/Classifications/OPS-ICHI/OPS/_node.html, last accessed: 31.01.2025

2 https://www.bfarm.de/DE/Kodiersysteme/Klassifikationen/ICD/ICD-10-GM/_node.html, last accessed: 31.01.2025

## Bibliography

[1] J. Styczyński et al., “Death after hematopoietic stem cell transplantation: changes over calendar year time, infections and associated factors.,” Bone Marrow Transplant., vol. 55, no. 1, pp. 126–136, Jan. 2020, doi: 10.1038/s41409-019-0624-z.

[2] M. L. Sorror et al., “Hematopoietic cell transplantation (HCT)-specific comorbidity index: a new tool for risk assessment before allogeneic HCT.,” Blood, vol. 106, no. 8, pp. 2912– 2919, Oct. 2005, doi: 10.1182/blood-2005-05-2004.

[3] T. Luft et al., “EASIX and mortality after allogeneic stem cell transplantation.,” Bone Marrow Transplant., vol. 55, no. 3, pp. 553–561, Mar. 2020, doi: 10.1038/s41409-019-0703-1.

[4] T. Jo et al., “A convolutional neural network-based model that predicts acute graft-versus-host disease after allogeneic hematopoietic stem cell transplantation.,” Commun Med (London), vol. 3, no. 1, p. 67, May 2023, doi: 10.1038/s43856-023-00299-5.

[5] R. Shouval et al., “Prediction of Allogeneic Hematopoietic Stem-Cell Transplantation Mortality 100 Days After Transplantation Using a Machine Learning Algorithm: A European Group for Blood and Marrow Transplantation Acute Leukemia Working Party Retrospective Data Mining Study.,” J. Clin. Oncol., vol. 33, no. 28, pp. 3144–3151, Oct. 2015, doi: 10.1200/JCO.2014.59.1339.

[6] Y. Arai et al., “Using a machine learning algorithm to predict acute graft-versus-host disease following allogeneic transplantation.,” Blood Adv., vol. 3, no. 22, pp. 3626–3634, Nov. 2019, doi: 10.1182/bloodadvances.2019000934.

[7] L. Lapp, M. Roper, K. Kavanagh, M.-M. Bouamrane, and S. Schraag, “Dynamic Prediction of Patient Outcomes in the Intensive Care Unit: A Scoping Review of the State-of-the-Art.,” J. Intensive Care Med., vol. 38, no. 7, pp. 575–591, Jul. 2023, doi: 10.1177/08850666231166349.

[8] H.-C. Thorsen-Meyer et al., “Dynamic and explainable machine learning prediction of mortality in patients in the intensive care unit: a retrospective study of high-frequency data in electronic patient records.,” Lancet Digit. Health, vol. 2, no. 4, pp. e179–e191, Apr. 2020, doi: 10.1016/S2589-7500(20)30018-2.

[9] G. P. Bodey, M. Buckley, Y. S. Sathe, and E. J. Freireich, “Quantitative relationships between circulating leukocytes and infection in patients with acute leukemia.,” Ann. Intern. Med., vol. 64, no. 2, pp. 328–340, Feb. 1966, doi: 10.7326/0003-4819-64-2-328.

[10] T. Prébet et al., “Platelet recovery and transfusion needs after reduced intensity conditioning allogeneic peripheral blood stem cell transplantation.,” Exp. Hematol., vol. 38, no. 1, pp. 55–60, Jan. 2010, doi: 10.1016/j.exphem.2009.10.004.

[11] S. G. Kong, S. Jeong, S. Lee, J.-Y. Jeong, D. J. Kim, and H. S. Lee, “Early transplantation-related mortality after allogeneic hematopoietic cell transplantation in patients with acute leukemia.,” BMC Cancer, vol. 21, no. 1, p. 177, Feb. 2021, doi: 10.1186/s12885-021-07897-3.

[12] F. Pedregosa et al., “Scikit-learn: Machine Learning in Python,” Journal of Machine Learning Research, vol. 12, no. 85, pp. 2825–2830, 2011.

[13] J. Van Der Donckt, J. Van Der Donckt, E. Deprost, and S. Van Hoecke, “tsflex: Flexible time series processing & feature extraction,” SoftwareX, vol. 17, p. 100971, Jan. 2022, doi: 10.1016/j.softx.2021.100971.

[14] F. Mölder et al., “Sustainable data analysis with Snakemake,” F1000Res., vol. 10, p. 33, Jan. 2021, doi: 10.12688/f1000research.29032.1.

