## Supplementary material for "Dynamic Prediction of Mortality Risk Following Allogeneic Hematopoietic Stem Cell Transplantation": Supplementary Note 1. Clinical Annotations.docx

### **Predominantly low risk, deceased**

- **B**: Passed away on day 44 in the ICU.
  - Shortly after transplantation: pneumonia and other infections.
  - Rapid deterioration; cause of death was kidney failure.
- **F**: Passed away on day 84.
  - Acute liver and kidney failure apparently not caused by bacterial or viral infection;
  - No prior indication of liver or kidney failure.
  - CRP increase in the first days after HSCT; later improvement.
  - Discharged in good general condition on day 21
  - Readmission on day 62.
  - Dialysis required.
- **G**: Passed away on day 84.
  - Discharge on day 2.
  - No medical report.
  - Readmission on day 48 with primary diagnosis T86.02 (aGvHD grade 3 and 4).
- **H**: Passed away on day 86.
  - Two short discharges (day 28–29; day 50–59), both in "good general condition."
  - Viral infection.
  - Pulmonary parenchymal hemorrhage as the cause of death.
  - Moderate risk on day 29 (readmission); fever, thrombocytopenia, no CRP increase.
  - D59.3: hemolytic-uremic syndrome.
  - ICU t
- **I**: Passed away on day 85 after readmission on day 78.
  - ICU t
  - Cardiovascular failure following pulmonary embolism (embolism appeared unexpectedly; extubated prior).
  - MDS patient.
  - Epstein-Barr virus detected.
  - Unclear space-occupying brain lesions.
  - Discharged post-transplantation on day 33 in good general condition.

### **Predominantly moderate risk, deceased**

- **C**: Passed away on day 13.
  - ITS Unit
  - Elevated infection markers shortly after transplantation without pathogen detection.
  - Tonic-clonic seizure.
  - Signs of pneumonia.
  - Passed away in poor general condition.
- **D**: Passed away on day 35.
  - Discharge day 19; readmission on day 28 due to vomiting, fever, diarrhea.
  - Outpatient radiotherapy post-transplantation (tumor regrowth near former lymphoma sites).
  - Therapy interruption before transplantation.
  - Cause of death: septic shock and multiorgan failure.

### **Predominantly high risk, deceased**

- **A**: Passed away on day 31; admission on day -36.
  - ICU
  - Day 5: atypical pneumonia and emergency gallbladder removal due to inflammation.
  - Cause of death: multiple organ failure.
- **E**: Passed away on day 69.
  - Discharge on day 11; readmission on day 18.
  - Discharged in good general condition.
  - Readmitted to ICU with Pneumocystis pneumonia.
  - Treatment caused kidney dysfunction.
  - Tracheal secretion positive for Klebsiella pneumoniae and Pseudomonas aeruginosa (both highly resistant).
  - Severe infections.

### **Predominantly high risk, alive**

- **K**: Transferred to another hospital on day 32.
  - Acute cutaneous and hepatic GvHD shortly after transplantation (improved with steroids).
  - Readmission on day 42 with fungal pneumonia.
  - Alive at least until day 296 (discharged in stable general condition).
  - Second discharge in good general condition but on parenteral nutrition.
  - Invasive pulmonary aspergillosis.

### **Predominantly low risk, alive**

- **J**: Discharged on day 14, no readmissions.
  - Always green.
  - Clinical course mostly uneventful, though with multiple bacterial and viral infections (well managed).
  - No relapse.
