## Supplementary material for "Dynamic Prediction of Mortality Risk Following Allogeneic Hematopoietic Stem Cell Transplantation": Supplementary Note 2. Patient ID Mappings.docx

### Misclassifications reflect clinical complexity

A 353

B 568

C 700

D 153

E 530

F 482

G 176

H 431

I 8

J 483

K 814

### High risk predictions can precede risk recognition by clinicians

10 Clinician would have detected earlier

82 Undecided

116 [X] Algorithm would have detected earlier

127 Algorithm would have detected earlier

153 Undecided

194 Algorithm would have detected earlier

220 Algorithm would have detected earlier

299 Clinician would have detected earlier

301 Clinician would have detected earlier

353 Clinician would have detected earlier

431 Algorithm would have detected earlier

502 Clinician would have detected earlier

530 Algorithm would have detected earlier

595 Algorithm would have detected earlier

618 Agreement

635 Algorithm would have detected earlier

665 [Y] Algorithm would have detected earlier

700 Algorithm would have detected earlier

772 Algorithm would have detected earlier
