## Supplementary material for "Dynamic Prediction of Mortality Risk Following Allogeneic Hematopoietic Stem Cell Transplantation": Supplementary Note 3. Assignment of ICD-10-GM Codes to HCT-CI Categories.docx

- **Arrhythmia:** I47 – I49
- **Heart disease:** I20 – I25, I50 – I52, I35 – I39
- **Inflammatory bowel diseases:** K50 – K51
- **Diabetes:** E10 – E14
- **Cerebrovascular diseases:** I63 – I64, G45
- **Psychiatric disorders:** F32 – F33, F40 – F41
- **Liver dysfunctions:** K73, K70.1, K71.3 – K71.5, K75.2, K75.3, B18; K74, K70.2, K70.3, K76.1, K71.7, P78.8
- **Obesity:** E66 \ {E66.x0, E66.x4, E66.x5}
- **Infections:** A00 – B89, B99
- **Rheumatic diseases:** M32; M05, M06; M33; M35.1; M35.3, M31.5
- **Gastric ulcer:** K25
- **Kidney dysfunctions:** Z49, Z99.2; Z94.0
- **Lung dysfunctions:** Z99.8, R06.0
- **Tumor:** C00 – C97 \ {C81 – C96, C44.9}; D00 – D48 \ {D45 – D47}

Exclusions are indicated with the notation " \ {...}".
