## Supplementary material for "Dynamic Prediction of Mortality Risk Following Allogeneic Hematopoietic Stem Cell Transplantation": Supplementary Note 4. Time Series Features.docx

**Minimum**

**Maximum**

**Mean**

**Median**

**Variance**

**Skewness**

**Quantiles**

(We calculate the quantiles for q = 0.25, 0.5, 0.75, 0.9, 0.95 or 0.99)

**Coefﬁcient of variation**

**Number of peaks (numerical); w = 1 or 2**

We divide a time series into overlapping subsequences of length 2*w+1. Whenever the middle point of a subsequence is the maximum of the subsequence this is counted as an observed peak.

**Number of peaks (continuous wavelet transform)**

We used the Ricker wavelet and varied the width between 1 and 6. The sum of the number of all peaks for all widths was used for this feature.

**Number of turning points (cubic spline interpolation)**

Based on a cubic spline interpolation the number of roots of the first derivative was used.

**Sample entropy**

We used the sample entropy as introduced by Richman and Moorman.

**Complexity-invariant distance**

We used the complexity-invariant distance as introduced by Batista et al. .

**Normalized CID**

To normalize we applied z-Score normalization.

**Autocorrelation (a = 3, 5, 7, 10 or 14)**

The variable a represents the position where the time series is split into two subseries and tested for autocorrelation.

**Percentage of recurring data points**

The fraction of non-unique values relative to the full time series

**Percentage of recurring values**

The fraction of non-unique values relative to all different, observed values
