## Supplementary figures and images for "Dynamic Prediction of Mortality Risk Following Allogeneic Hematopoietic Stem Cell Transplantation"

### Supplementary Figure 1. Patient Trajectories.png

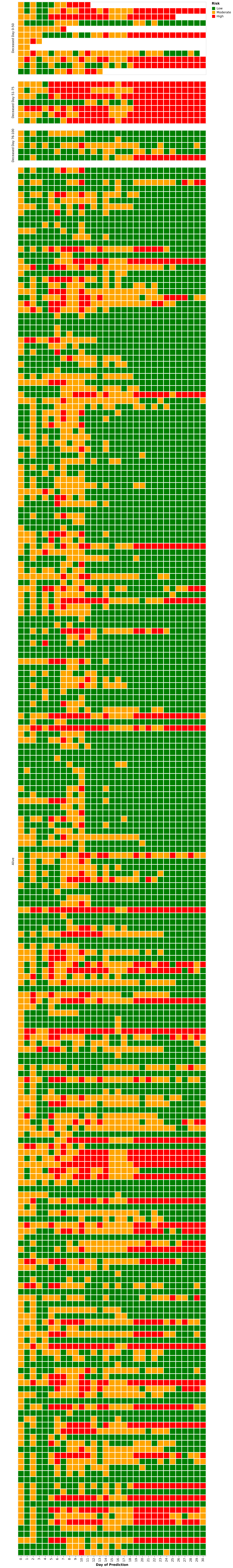

### Supplementary Figure 2. Feature Recency.png

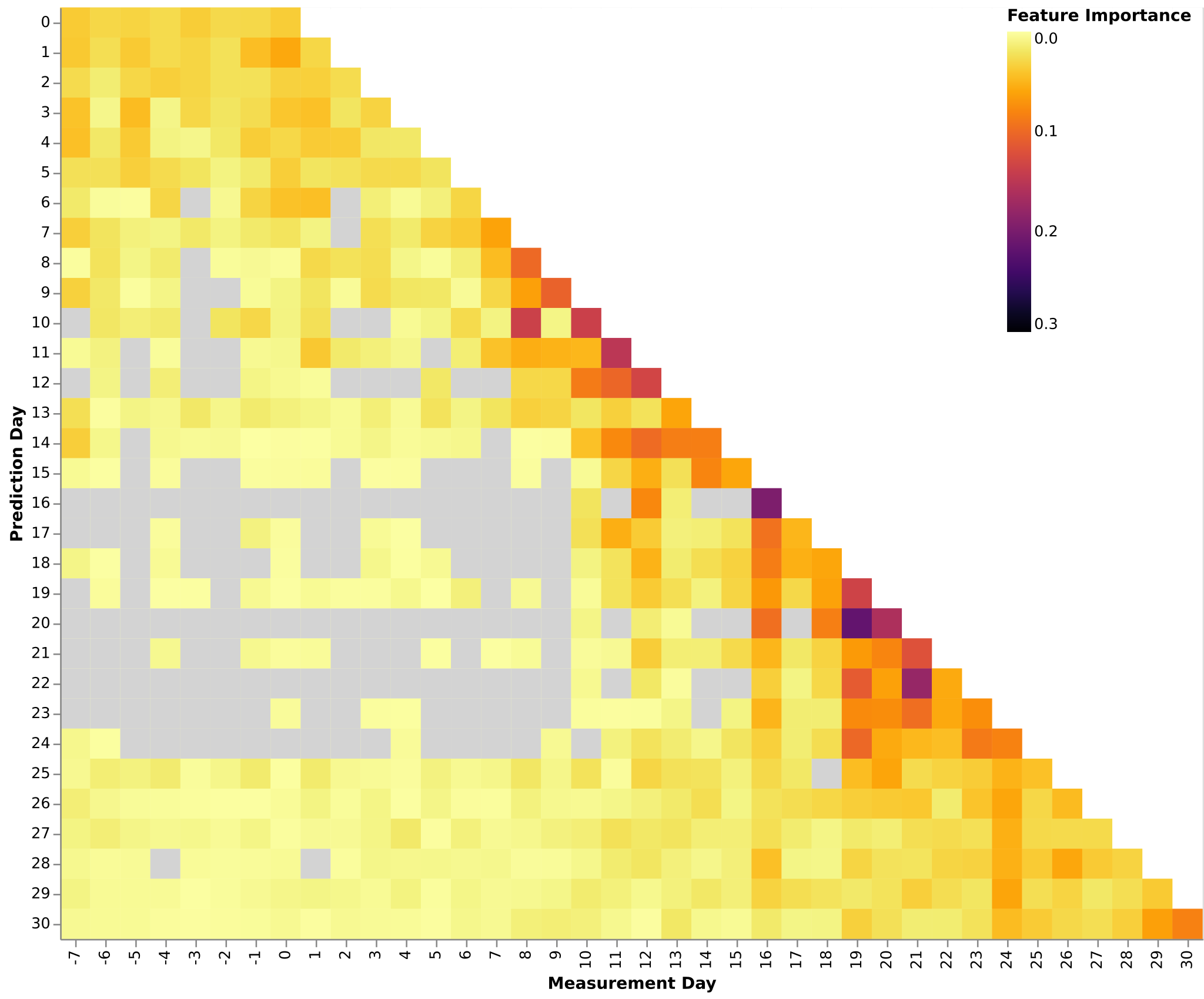
